# Comparison of Accelerometry-based Measures of Physical Activity

**DOI:** 10.1101/2022.03.16.22272518

**Authors:** Marta Karas, John Muschelli, Andrew Leroux, Jacek K Urbanek, Amal A Wanigatunga, Jiawei Bai, Ciprian M Crainiceanu, Jennifer A Schrack

## Abstract

**Background:** Given the evolution of processing and analyzing accelerometry data over the past decade, it is of utmost importance that we as a field understand how newer (e.g., MIMS) summary measures compare to long-established ones (e.g., ActiGraph activity counts).

**Objective:** Our study aims to compare and harmonize accelerometry-based measures of physical activity (PA) to increase the comparability, generalizability, and translation of findings across studies using objective measures of PA.

**Methods:** High resolution accelerometry data were collected from 655 participants in the Baltimore Longitudinal Study on Aging who wore an ActiGraph GT9X device at wrist continuously for a week. Data were summarized at the minute-level as activity counts (AC; measure obtained from ActiGraph’s ActiLife software) and MIMS, ENMO, MAD, and AI (open-source measures implemented in R). The correlation between AC and other measures was quantified both marginally and conditionally on age, sex and BMI. Next, each pair of measures were harmonized using nonparametric regression of minute-level measurements.

**Results:** The study sample had the following characteristics: mean (sd) age of 69.8 (14.2), BMI of 27.3 (5.0) kg/m^2^, 54.5% females, and 67.9% white. The marginal participant-specific correlation between AC and MIMS, ENMO, MAD, and AI were 0.988, 0.867, 0.913 and 0.970, respectively. After harmonization, the mean absolute percentage error for predicting total AC from MIMS, ENMO, MAD, and AI was 2.5, 14.3, 11.3 and 6.3, respectively. The accuracy for predicting sedentary minutes based on AC (AC > 1853) using MIMS, ENMO, MAD and AI was 0.981, 0.928, 0.904, and 0.960, respectively. An R software with a unified interface for computation of the open-source measures from raw accelerometry data was developed and published as SummarizedActigraphy R package.

**Conclusions:** Our comparison of accelerometry-based measures of PA enables researchers to extend the knowledge from the thousands of manuscripts that have been published using ActiGraph AC to MIMS and other measures by demonstrating their high correlation and providing a harmonization mapping.

## Introduction

Accelerometer-based activity monitors have become increasingly popular in research studies because they provide non-invasive, objective measures of physical activity (PA) that can be collected continuously for extended periods of time (Karas et al., 2019). Modern wearable accelerometers measure acceleration of a body at high frequency (typically 10-100 Hz). These raw data are then typically aggregated into fixed-time epochs, e.g. 1 minute-long. Yet, the choice of epoch-based measures varies across studies. For example, the Baltimore Longitudinal Study on Aging (BLSA) used wrist-worn accelerometers and summarized data using activity counts (AC) (Neishabouri et al., 2022), a measure proposed and implemented by ActiGraph (ActiGraph LLC, Pensacola, FL, USA). The wrist-worn accelerometry data collection in the recent NHANES 2011-2014 opted for Monitor-Independent Movement Summary (MIMS) (John et al., 2019), an open-source summary characteristic of high-density accelerometry data. The UK Biobank study used wrist-worn accelerometers and chose Euclidean Norm Minus One (ENMO) (van Hees et al., 2013), a different open-source summary measure of high-density accelerometry data. Additional open-source summary measures of acceleration are Mean Amplitude Deviation (MAD) (Vähä-Ypyä et al., 2015) and Activity Intensity (AI) (Bai et al., 2014).

Given the evolution of processing and analyzing accelerometry data over the past decade, it is of utmost importance that we as a field understand how newer (e.g., MIMS) summary measures compare to long-established ones (e.g., ActiGraph AC). Further, we recognize the need to harmonize, or map, values of PA summaries derived from different algorithms. This enables us to extend the knowledge from the thousands of manuscripts that have been published using ActiGraph AC (where no repository or access to raw accelerometry data remains available) to MIMS and other measures.

To address this problem, high resolution accelerometry data were examined from 655 participants in the Baltimore Longitudinal Study on Aging (BLSA) who wore an ActiGraph GT9X Link device at wrist continuously for a week. Data were summarized in 1-minute epochs as ActiGraph AC (obtained from ActiLife Software), MIMS, ENMO, MAD, and AI (open-source measures implemented in R). The correlations between ActiGraph AC and MIMS, ENMO, MAD, and AI were quantified both marginally and conditionally on age, sex, and body mass index (BMI). This provides simple summaries of associations and a guide for the strength of these associations in subgroups defined by demographic information. Next, ActiGraph AC was harmonized with each of the other measures using nonparametric smoothing of minute-level data. This allows to: (1) provide a mapping between any two PA summary measures considered; (2) derive cut-points of open-source PA measures that correspond to established cut-points to estimate time spent in different PA intensities for ActiGraph AC. Our analysis is especially timely given the recent release of physical activity data from NHANES 2011-2014 that uses the open-source MIMS measure. In the remaining part of the manuscript, “AC” alone is used to denote ActiGraph AC.

## Methods

### Study Design and Population

Data used in this manuscript were collected as part of the National Institute on Aging’s Baltimore Longitudinal Study of Aging (BLSA). The study has been active since 1958 with the aim to describe longitudinal physical and cognitive changes related to aging. Participant enrollment criteria and a general description of the sample have been reported elsewhere (Kuo et al., 2020). Briefly, participants are community dwelling volunteers free of all major chronic conditions and cognitive and functional impairment at the time of enrollment. Participants undergo a comprehensive health and functional screening evaluation at baseline and are followed for life, attending follow-up visits and extensive health testing every 1-4 years depending on age. The study protocol was approved by the Internal Review Board of the Intramural Research Program of the National Institutes of Health. The data used in this work were collected in all participants who agreed to wear an accelerometer between July 2015 and January 2019.

### Accelerometry Data Collection and Export

Data were collected with an ActiGraph GT9X Link device, a tri-axial accelerometer that measures time-varying body accelerations in magnitudes ranging ± 8 *g* (*g* = 9.81 m/s^2^). The devices were configured to record data at a frequency of 80 Hz. Participants were given the monitor on the last day of their clinic visit and were instructed to wear it at all times on their non-dominant wrist for 7 days, except for periods of extended swimming or bathing. Devices were returned to the clinic via a pre-addressed envelope. The ActiLife software (version 6.13.4) was used to: (a) export data into the “gt3x” file format ; (b) derive minute-level AC and export them as CSV files; and (c) export raw acceleration data as three-dimensional time series in *g* units together with subsecond-level timestamps into CSV files. In AC derivation, the ActiGraph’s low-frequency extension was used following the recommendation from Cain et al. (2013).

### Raw Accelerometry Data Quality Control

Three raw data quality check flags were adapted from a set of nine flags recently introduced by the release of the NHANES 2011-2012 wave protocol (NHANES 2011-2012 Data Documentation, 2020). The selected flags subset represents intuitive flags that are meant to “determine signal patterns that were unlikely to be a result of human movement” but are not aimed at identifying non-wear. The flags definitions are given in Appendix A. A raw data observation was flagged as valid if it had none of the three flags triggered; otherwise, it was marked as invalid. Raw data observation flags were further used to mark data for exclusion at minute-level (see Sect. “Minute-level Accelerometry Data Preprocessing” below).

### Open-source Summary Measures of Raw Accelerometry Data

The raw accelerometry data were used to derive a set of commonly used minute-level open-source summary measures: MIMS, ENMO, MAD, and AI. Definitions of the measures are specified in Appendix B. The calculate_measures method from the SummarizedActigraphy R package (Muschelli, 2021) was used to compute the measures. The method’s primary contribution is to provide a unified data interface to compute a range of open-source measures; it uses references to some of the measures’ original software, e.g. the MIMSunit R package (Tang et al., 2020) to compute MIMS.

### Minute-level Accelerometry Data Preprocessing

To define minute-level sensor wear/non-wear flags, the get_wear_flag method from the arctools R package (Karas et al., 2021) was used. The method implements a wear/non-wear detection algorithm proposed in Choi et al. (2011); it uses threshold of 0 for the number of nonzero counts allowed during a non-wear time interval, and it does not allow “artifactual movement” interval of nonzero counts during a non-wear time interval. A recommended value of 90 minutes was used for the minimum time of consecutive zero counts for a window to be flagged as non-wear. A minute had a valid raw data flag if no quality control flags at raw observation-level were triggered within that minute (see Sect. “Raw Accelerometry Data Quality Control” above). Finally, a minute was defined as a valid minute (overall) if it had both a wear flag and valid raw data flag positive; otherwise, it was defined as an invalid minute. A valid day was defined as a day (12:00am-11:59pm) with no more than 10% (144 minutes) invalid minutes (Wanigatunga et al., 2020). Only participants who had at least 3 valid days of data, and only their valid days data, were included in further preprocessing and analyses.

Values of AC, MIMS, ENMO, MAD and AI were winsorized (Hastings Jr. et al., 1947) by first computing a measure-specific 0.999 quantile, and then using it to replace the measure’s values exceeding the quantile’s value. As a result, 0.1% of each measure’s most extreme values were replaced in the dataset. The winsorization reduces the effect of extreme values.

Lastly, a separate data set was constructed where values of AC, MIMS, ENMO, MAD and AI were imputed for invalid minutes, following the imputation approach previously used in Leroux et al. (2020). This data set was used in the summary of daily sums of measure values and in our application example where data without missing values were needed. The imputation procedure was conducted separately for each measure. The procedure started by computing a functional principal component analysis (FPCA) of functional observations made of all participant- and day-specific minute-level data parts, each sorted by time and arranged into a 1440-long vector. A fast implementation of the sandwich smoother (Xiao et al., 2013) from fpca.face method in the refund R package (Goldsmith et al., 2020) was used to allow for a quick FPCA computation given our dataset volume. The smoothed version of each functional observation was obtained from FPCA, and their values were used for data imputation.

### Statistical Data Analysis

A summary of minute-level measures: AC, MIMS, ENMO, MAD, and AI was computed in two different ways: (a) as average daily sum per participant, summarized across participants; (b) as average per minute, summarized across all participant-minutes.

A Pearson correlation coefficient between four pairs of measures: AC and MIMS, AC and ENMO, AC and MAD, and AC and AI, was computed for each participant. Mean correlation values and its standard error were quantified via intercept-only linear regression models in which participant-specific correlation between a pair of measures was set as an outcome. Next, the demographics effect on correlation was estimated via adjusted linear regression models. Participant-specific correlation between a pair of measures was set as an outcome, and age, BMI and sex (is male) indicator were set as covariates. Significance level alpha = 0.05 was assumed in determining statistical significance of coefficients in the adjusted models. For both unadjusted and adjusted models, a separate fit was estimated for each four pairs of measures.

To derive the harmonization mapping, relationships between measures: AC and MIMS, AC and ENMO, AC and MAD, and AC and AI, were estimated with generalized additive models (GAMs) separately for each pair of measures. The GAMs were chosen to allow flexible adaptation to the data rather than imposing a particular functional form of the fit. In each model, the outcome consisted of minute-level open-source measure, and predictors were a smooth term of minute-level AC. For the smooth term, cubic regression splines with a basis dimension equal to 30 were used to allow a flexible relationship between a measure and AC. Models were estimated with nonparametric smoothing implemented in gam method in the mgcv R package (Wood, 2021). Smoothness of the non-linear effects was enforced via a second derivative penalty with smoothing parameter selection done using cross-validation (Wood, 2011). Data from all participants’ valid minutes were used in the model fitting except minutes which had AC values equal 0. The AC = 0 exclusion was motivated by a large proportion of zero values, and the need to estimate the relation for small AC values without it being inflated by the large number of zeros. All four relationships were estimated as strictly monotonic (without monotonicity having been constrained explicitly). The GAM model-fitted values were used to define two-way mappings between AC and each of the four open-source measures. All open-source measurements were mapped into AC. For the remainder of the manuscript, we denote 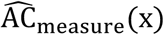 to be AC value estimated via the mapping from x value of a measure, where “measure” stands for one of: MIMS, ENMO, MAD, AI.

Several steps were taken to assess the mapping accuracy. First, to assess mapping accuracy in estimating PA volume statistics, total activity count (TAC; the sum of minute-level values from a day) was computed for each participant’s day, using both true AC and 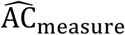. The participant- and day-specific estimation error was defined as the difference between TAC derived from true AC and TAC derived from 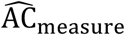. Participant-specific mean percentage error (MPE) and mean absolute percentage error (MAPE) were computed by averaging the error across participant’s days. Second, to assess whether mapping accuracy depends on participant’s activity level, participant-specific MPE were plotted against participant’s average TAC (similar to a Bland-Altman plot). Third, the mapping utility in classifying minutes into sedentary versus active was assessed. In the classification task, for each minute, the minute’s label was defined based on whether AC > 1853 (Koster et al., 2016) using true AC value, and the prediction value was defined based on whether 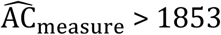. Accuracy, sensitivity and specificity of prediction were computed for each participant.

Minute-level AC and 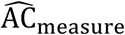 were used to estimate smoothed 24-hour median activity counts across the previously published (Schrack et al., 2013) age groups: < 60-year old, 60-to 67-year old, 68- to 74-year old, ≥ 75-year old. The similarity between AC-based and 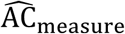-based estimates were summarized using MAPE (sum of absolute value of difference between estimates divided by sum of AC-based estimates) separately for each measure: MIMS, ENMO, MAD, AI.

The project repository containing all code for data preprocessing and analysis is publicly available on GitHub (muschellij2/blsa_mims).

## Results

### Population Characteristics

The final study sample consisted of N=655 individuals whose characteristics are detailed in Table 1. The mean age was 69.8 (SD = 14.2, range 22-97) years. There was a higher proportion of women (54.5%) compared to men (45.5%). The racial composition reflected that of the BLSA enrollment: 68% white, 24% black, 7% other (1% not reported). Almost 96% of participants self-reported good, very good or excellent health. The prevalence of hypertension, high blood cholesterol, and osteoarthritis was 44%, 53%, and 48%, respectively. All other chronic conditions were rare with a prevalence of < 15%. Participants had a median 6 (range 3-7) days of valid accelerometry data; within these, they had an average of 1438 (SD = 8) out of 1440 valid minutes per day.

**Table 1.** Study sample (N = 655) characteristics.

|  | Mean (SD) | Median [min, max] | n (% of N) |
| --- | --- | --- | --- |
| Age | 69.8 (14.2) | 72.0 [22.0, 97.0] |  |
| Weight [kg] | 77.4 (17.1) | 76.3 [41.1, 142.7] |  |
| Height [cm] | 168.0 (9.2) | 167.3 [143.8, 196.2] |  |
| BMI | 27.3 (5.0) | 26.6 [17.1, 52.5] |  |
| **Sensor wear** |  |  |  |
| Valid days | 5.9 (0.4) | 6.0 [3.0, 7.0] |  |
| Non-wear min./day | 2.0 (7.8) | 0.0 [0.0, 77.0] |  |
| Valid min./day | 1437.8 (8.0) | 1440.0 [1361.7, 1440.0] |  |
| **Sex** |  |  |  |
| Female |  |  | 357 (54.5) |
| Male |  |  | 298 (45.5) |
| **Race** |  |  |  |
| White |  |  | 445 (67.9) |
| Black |  |  | 157 (24.0) |
| Other |  |  | 44 (6.7) |
| Not reported |  |  | 9 (1.4) |
| **Self-rep. health<sup>a</sup>** |  |  |  |
| Gd/vgd/excell<sup>b</sup> |  |  | 628 (95.9) |
| Fair/poor |  |  | 22 (3.4) |
| Not reported |  |  | 5 (0.8) |
| **Medical history** |  |  |  |
| MI/CHF/A/VP/PAD<sup>c</sup> |  |  | 55 (8.4) |
| Hypertension |  |  | 285 (43.5) |
| High blood chol. <sup>d</sup> |  |  | 346 (52.8) |
| Stroke/TIA <sup>e</sup> |  |  | 34 (5.2) |
| Pulmonary disease |  |  | 74 (11.3) |
| Diabetes |  |  | 95 (14.5) |
| Cancer |  |  | 191 (29.2) |
| Osteoarthritis |  |  | 316 (48.2) |

<sup>a</sup>Self-rep. health = Self-reported health.
<sup>b</sup>Gd/vgd/excell = Good/very good/excellent.
<sup>c</sup>MI/CHF/A/VP/PAD = Heart attack/congestive heart failure/ischemic chest pain/vascular procedure/peripheral artery disease.
<sup>d</sup>High blood chol. = High blood cholesterol.
<sup>e</sup>TIA = Transient ischemic attack.

Table 2 describes minute-level measures: AC, MIMS, ENMO, MAD and AI, computed as an average daily sum per participant, and then summarized across participants. Daily AC had mean (sd) 2,204,169 (600,965), and MIMS, MAD, ENMO and AI had average (sd) 11,299.7 (2766.0), 47.7 (13.3), 30.9 (9.1) and 4157.6 (1068.8), respectively. Table C1 in Appendix C describes minute-level measures summarized as average per minute, across all participant-minutes.

**Table 2.** Summary of average daily sum of AC, MIMS, ENMO, MAD, and AI. Each value in the table is an aggregate -- mean, standard deviation, median, minimum and maximum -- of N = 655 participant-specific values of average daily sum. The summary was computed based on a data set of winsorized, invalid minutes-imputed measures.

|  | Mean (SD) | Median [min, max] |
| --- | --- | --- |

| Measure name |  |  |
| --- | --- | --- |
| AC | 2204169 (600965) | 2157496 [731945, 5071196] |
| MIMS | 11299.7 (2766.0) | 11195.2 [4252.3, 23931.5] |
| MAD | 47.7 (13.3) | 46.3 [16.1, 108.1] |
| ENMO | 30.9 (9.1) | 29.6 [11.8, 75.3] |
| AI | 4157.6 (1068.8) | 4085.5 [1529.7, 9418.6] |

### Correlations Between Minute-level Summary Statistics

Table 3 summarizes participant-specific correlation for pairs of minute-level measures: AC and MIMS, AC and ENMO, AC and MAD, AC and AI. Column 2 (“Model unadjusted”) shows estimated intercept coefficient and its standard error from an unadjusted (intercept-only) regression model. Marginally, the AC measure had the highest correlation with MIMS -- estimated mean (SE) 0.988 (0.0002), closely followed by that of AI -- 0.970 (0.0007). Correlation between AC and MAD had mean (SE) 0.913 (0.0013), and correlation between AC and ENMO -- mean (SE) 0.867 (0.0018).

**Table 3.**
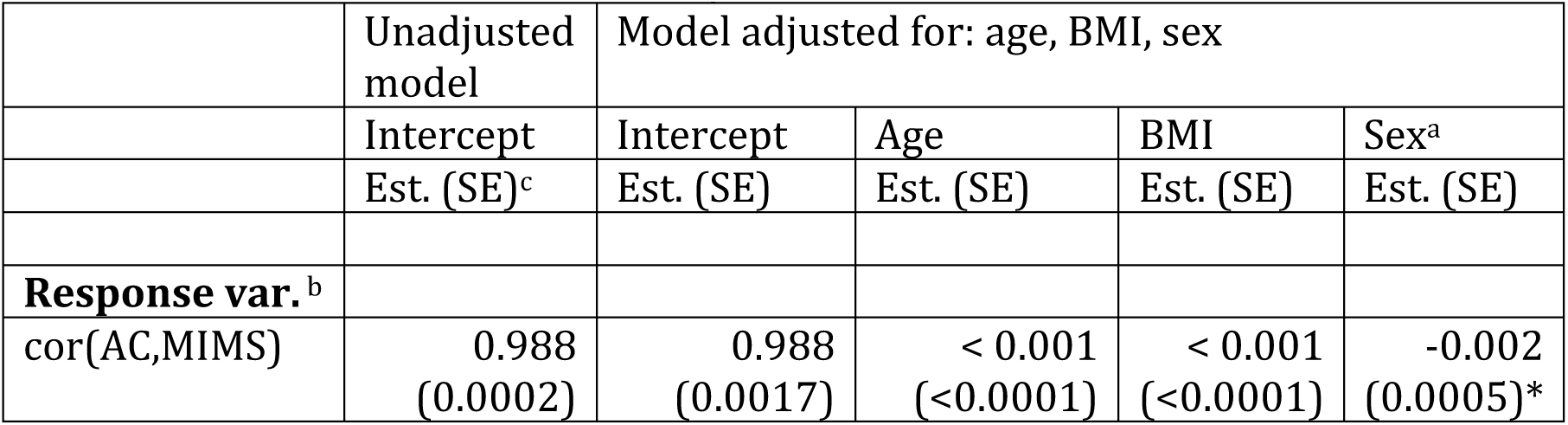
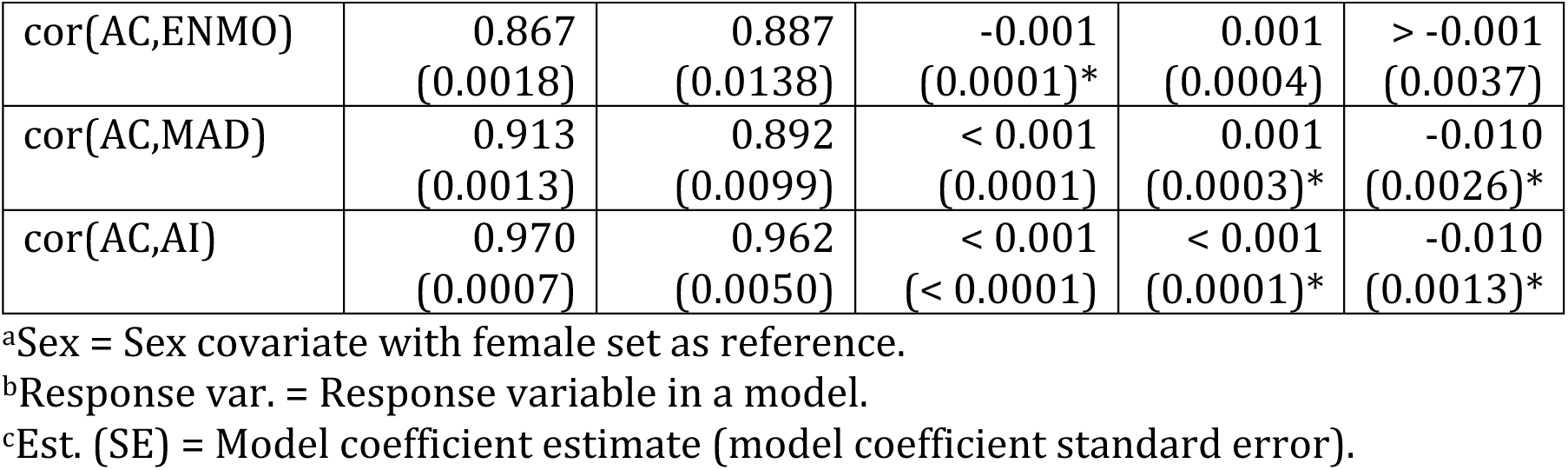
Summary of Pearson correlation for pairs of minute-level measures: AC, MIMS, ENMO, MAD, AI. In all models, participant-specific value of correlation was set as an outcome. The “*” symbol is used to denote model coefficients (excluding intercept) for which the corresponding p-value was <0.05.

|  | Unadjusted model | Model adjusted for: age, BMI, sex |  |  |  |
| --- | --- | --- | --- | --- | --- |
|  | Intercept | Intercept | Age | BMI | Sex<sup>a</sup> |
|  | Est. (SE)<sup>c</sup> | Est. (SE) | Est. (SE) | Est. (SE) | Est. (SE) |
| Response var.<sup>b</sup> |  |  |  |  |  |
| cor(AC,MIMS) | 0.988<br>(0.0002) | 0.988<br>(0.0017) | < 0.001<br>(<0.0001) | < 0.001<br>(<0.0001) | -0.002<br>(0.0005)* |
| cor(AC,ENMO) | 0.867<br>(0.0018) | 0.887<br>(0.0138) | -0.001<br>(0.0001)* | 0.001<br>(0.0004) | > -0.001<br>(0.0037) |
| cor(AC,MAD) | 0.913<br>(0.0013) | 0.892<br>(0.0099) | < 0.001<br>(0.0001) | 0.001<br>(0.0003)* | -0.010<br>(0.0026)* |
| cor(AC,AI) | 0.970<br>(0.0007) | 0.962<br>(0.0050) | < 0.001<br>(< 0.0001) | < 0.001<br>(0.0001)* | -0.010<br>(0.0013)* |

<sup>a</sup>Sex = Sex covariate with female set as reference.
<sup>b</sup>Response var. = Response variable in a model.
<sup>c</sup>Est. (SE) = Model coefficient estimate (model coefficient standard error).

Columns 3-6 (“Model adjusted”) show coefficient estimates and standard errors from a conditional model where participant-specific correlations were quantified while adjusting for age, BMI and sex. The estimated effect of age on the correlation was not statistically significant (alpha = 0.05) and had point estimates of magnitude less than 0.001 in models except the correlation between AC and ENMO (est. = -0.001, SE = 0.0001, p-value < 0.0001). The estimated effect of BMI on the correlation was statistically significant in models for correlation between AC and MAD (est. = 0.001, SE = 0.0003, p-value = 0.0013) and AC and AI (est. < 0.001, SE = 0.0001, p-value = 0.0362). The estimated effect of male sex (compared to female -- reference level) was statistically significant in three models: model of correlation between AC and MIMS (est. = -0.002, SE = 0.0005, p-value < 0.0001), AC and MAD (est. = -0.01, SE = 0.0026, p-value < 0.0001), AC and AI (est. = -0.01, SE = 0.0013, p-value < 0.0001).

### Mapping Between Minute-level Summary Measures

Figure 1 shows estimated association between minute-level AC (x-axis) and minute-level MIMS, ENMO, MAD, and AI, respectively (y-axis). The black solid line shows GAM model-fitted values of each measure (MIMS, ENMO, MAD, AI) given the AC measure value. Here, the GAM estimates can be thought of as a smoothed mean across the points.

**Figure 1.**
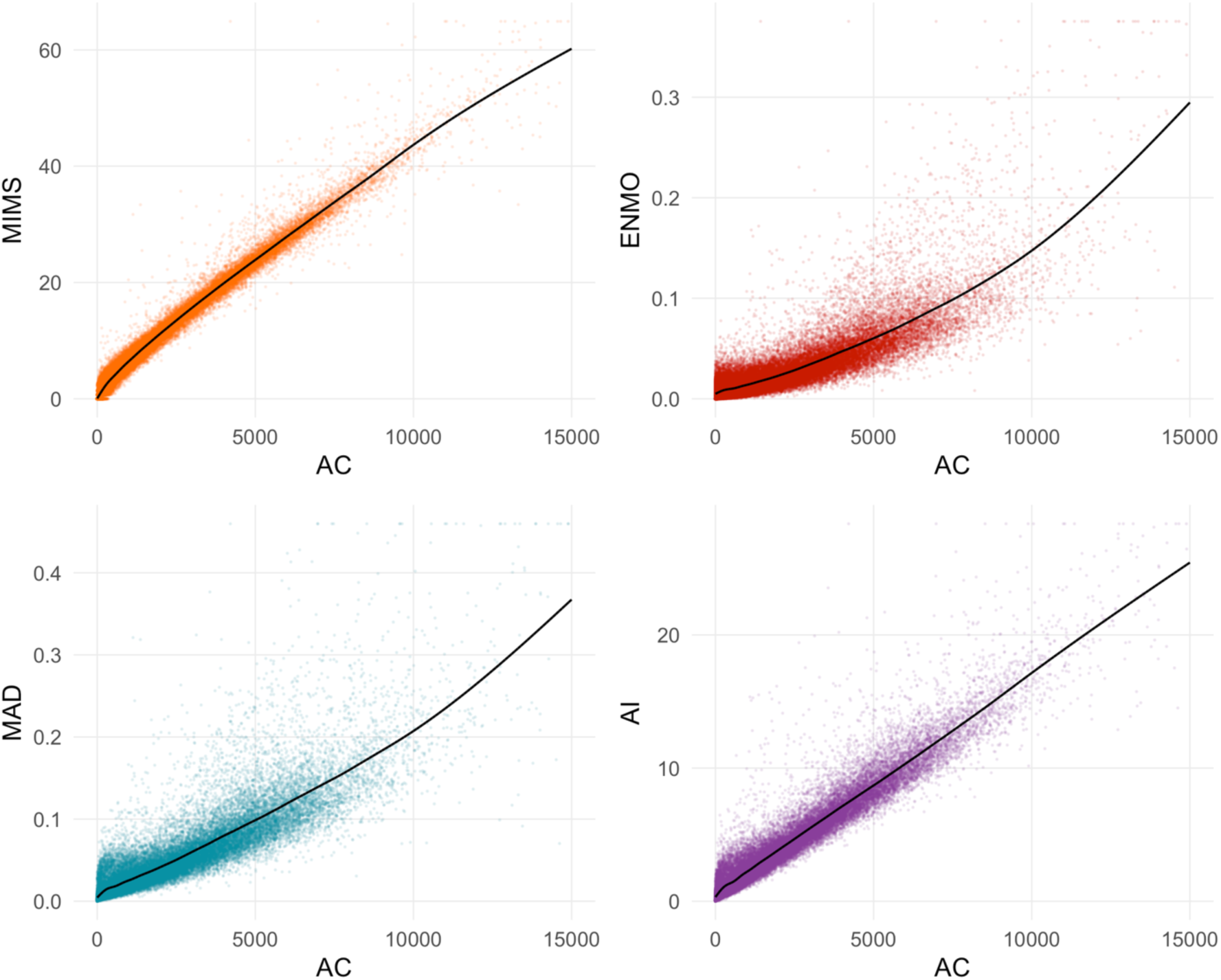
Estimated mapping between minute-level AC (x-axis) and minute-level MIMS, ENMO, MAD, and AI, respectively (y-axis). Black solid line shows GAM model-fitted values of a measure (MIMS, ENMO, MAD, AI) given the AC measure value. The points represent a subset of the data created by taking every 100-th observation from all participant- and minute-specific observations; this subset is the same across the four plots.

The CSV table with model-fitted values of MIMS, ENMO, MAD, and AI is publicly available on GitHub (muschellij2/blsa_mims). The results were used to implement fast R software that maps values of measure A to measure B that is also available on GitHub. Table 4 shows model-fitted values for selected published cut-off values of AC. Cut-off AC = 1853 was derived by Koster et al. (2016) to separate sedentary and active minutes from non-dominant wrist-worn sensor data collected in older adults. Cut-offs AC = 2860 and 3940 were derived by Montoye et al. (2020) to separate sedentary, light, and moderate-to-vigorous activity intensity levels from non-dominant wrist-worn sensor data collected in young to older adults. For a widely used cut-off AC = 1853, the fitted values were: MIMS = 10.558, ENMO = 0.022, MAD = 0.039, AI = 3.620.

**Table 4.** AC and fitted values of MIMS, ENMO, MAD and AI, respectively, for selected AC cut-off values.

|  | MIMS fitted | ENMO fitted | MAD fitted | AI fitted |
| --- | --- | --- | --- | --- |
| **AC cut-off** |  |  |  |  |
| 1853 | 10.558 | 0.022 | 0.039 | 3.620 |
| 2860 | 15.047 | 0.033 | 0.057 | 5.273 |
| 3940 | 19.614 | 0.046 | 0.078 | 7.025 |

The mapping accuracy in estimating PA volume statistics was quantified by participant-specific mean percentage error (MPE) and mean absolute percentage error (MAPE) in estimating total activity count (TAC). Table C2 in Appendix C summarizes the MPE and MAPE across the participants. The MAPE was the smallest for MIMS and had mean (SD) of 2.5 (2.4), followed by that of AI (mean = 6.3, SD = 5.1), MAD (mean = 11.3, SD = 8.4) and ENMO (mean = 14.3, SD = 10.3). The MPE were equal for MIMS -- mean (SD) 0.2 (3.2), for AI -- 0.3 (7.6), for MAD -- (−0.3) (13.3), and for ENMO -- 4.6 (16.1). Figure C1 in Appendix C shows participant-specific MPE arranged according to the participant’s average TAC. Based on visual inspection, there is a larger variability of MPE values among participants with smaller average TAC values, but there is no apparent tendency for lower or higher MPE values depending on participant’s average TAC.

The mapping utility in the task of classifying minutes into sedentary versus active using the AC = 1853 cut-off was also assessed. Table C3 in Appendix C summarizes participant-specific accuracy, sensitivity and specificity of predicting whether a minute is active, where the minute’s label was based on true AC and the prediction was based on 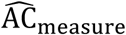. The participant-specific classification accuracy was the highest for MIMS and had mean (SD) of 0.981 (0.005), followed by that of AI (mean = 0.960, SD = 0.012), MAD (mean = 0.928, SD = 0.021) and ENMO (mean = 0.904, SD = 0.028).

### Minute-level Patterns of Daily Physical Activity

Previous work in the BLSA characterized age-related differences in daily patterns of physical activity using minute-level counts from Actiheart activity monitor (Schrack et al., 2013). Figure 2 shows the estimated smoothed 24-hour median activity counts across the previously published age groups: < 60-year old (green; N = 140), 60- to 67-year old (red; N = 102), 68- to 74-year old (blue; N = 129), ≥ 75-year old (orange; N = 284). Solid semi-transparent colour lines represent results obtained with AC measure. Dashed colour lines represent results obtained with 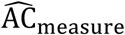 values mapped into AC from one of the four measures -- MIMS, ENMO, MAD, and AI -- per plot. Based on visual inspection, in each case, the 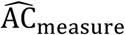-based curves yielded roughly the same information as the AC-based curves. The similarity between AC-based and 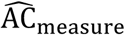-based estimates was summarized using mean absolute percentage error (MAPE). The measure-specific MAPE was the smallest for MIMS and equaled 3.2, followed by that of AI (6.7), MAD (11.1) and ENMO (12.5).

**Figure 2.**
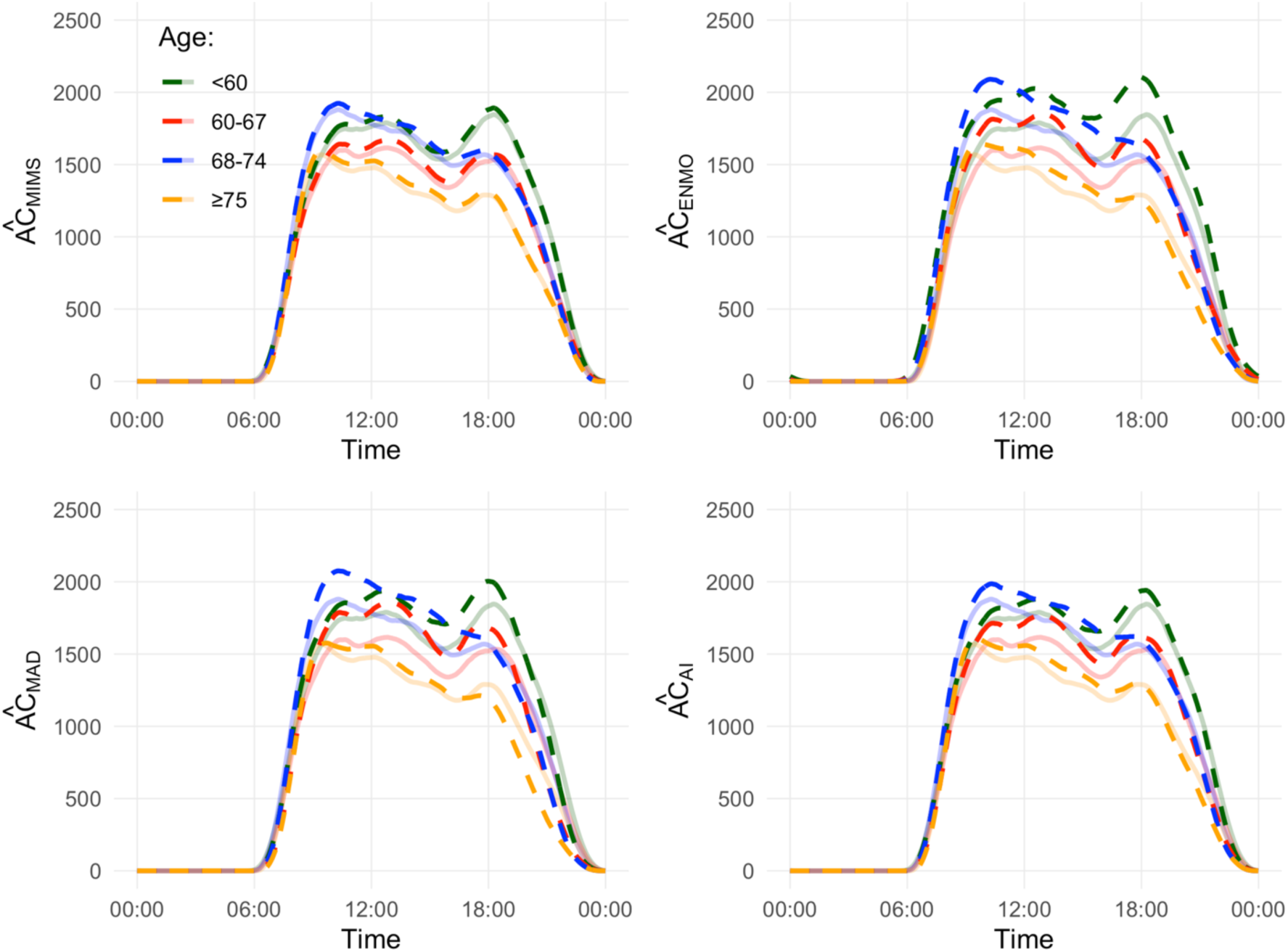
Smoothed 24-hour median activity counts per minute for each age group: < 60-year old (green), 60- to 67-year old (red), 68- to 74-year old (blue), ≥ 75-year old (orange). Solid semi-transparent colour lines represent results obtained with AC measure; they are the same across the four plots. Dashed colour lines represent results obtained with 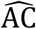 values mapped into AC from one of the four measures -- MIMS, ENMO, MAD, and AI -- per plot.

## Discussion

### Principal Results

We used a large-size cohort of BLSA participants (age mean 69.8, range 22-97) to compute and compare minute-level accelerometry-derived measures of physical activity: AC and MIMS, ENMO, MAD, and AI. Results suggest that correlations between the widely published AC and the other raw data summary metrics are all large (>= 0.87), and especially high for MIMS and AI (>= 0.97). After the harmonization, MIMS allowed for excellent accuracy in predicting TAC and predicting sedentary minutes with AC = 1853 cut-off. The observed differences in the correlations with AC between open-source measures should be considered when comparing historical results across the metrics.

Previously, the correlation between AC and MIMS in data collected during continuous monitoring in the free-living environment has not been explored. We computed participant-specific correlations between AC and MIMS, ENMO, MAD, and AI measures, and examined how the correlations differ across age, BMI and sex. The AC measure had the highest average participant-specific correlation with MIMS -- 0.988, closely followed by AI -- 0.97, and MAD (0.913) and ENMO (0.867). Both MIMS and AI are based on variability within each axis, whereas MAD and ENMO are based on the Euclidean norm of three-dimensional time-series of the raw data. Therefore, it is consistent with expectations to observe MIMS and AI behave similarly, and demonstrate similar correlations with AC. While we found statistically significant effects of age, BMI and sex on the correlations between AC and the other measures, the effect sizes were of very small magnitude. In particular, the analysis showed that MIMS had a correlation with AC that did not differ significantly across age nor BMI, and differed significantly between men and women by a magnitude of 0.002.

We estimated (and provided software to use) the harmonization mapping between minute-level measures -- AC and MIMS, AC and ENMO, AC and MAD, and AC and AI. The mapping allows us to extend the knowledge from the thousands of manuscripts that have been published using AC to MIMS and other measures in cases where the access to raw accelerometry data from a published work is no longer available. The mapping can be particularly useful to translate commonly used cut-off values of PA intensity levels from AC into open-source measures. The mapping was validated in the tasks of predicting TAC and predicting sedentary minutes based on AC = 1853 cut-off; we observed excellent accuracy for MIMS and AI of 0.981 and 0.96, respectively. The utility of the derived mapping was demonstrated in the example in which previous BLSA results were replicated. The PA volume daily trajectories across four age groups obtained with AC were closely matched with open-source measures, with MIMS yielding visually almost identical results (MAPE = 3.2), followed by that of AI, MAD and ENMO (MAPE = 6.7, 11.1, and 12.5).

Lastly, we believe we are the first to provide freely available R software (SummarizedActigraphy R package) with a unified interface for computation of the four open-source measures from raw accelerometry data. This effort is accompanied by the Appendix material where we provide a description of the used PA measures that distills complicated mathematical formulas into a reader-friendly text.

### Limitations

We identify the following limitations of our work. First, our study sample consists of predominantly middle-aged to older adults; specifically, < 30% of the sample is younger than 64 years and no children or adolescents below 22 years of age were included. However, we observed that: (a) the level of activity of adults in our study sample did range from sedentary to moderate and vigorous activity, (b) our mapping results did not exhibit any trend depending on average level of participant’s PA, and (c) the measured variability along the estimated mapping is lower for higher activity values, which suggests the derived mapping could prove similarly useful in future studies including younger (more active) populations. Second, PA measures were computed using raw accelerometry data collected at frequency 80 Hz; while this frequency matches the frequency of raw accelerometry data collection in recent release of PA data from NHANES 2011-2014 that uses the MIMS measure, studies collecting raw data of different frequency should use caution in adapting our harmonization mapping. Third, our comparison is limited to data collected with a sensor worn on a non-dominant wrist. While we expect the results to translate to a dominant-wrist, we presume the correlations and mapping may not be applicable to e.g. chest- or hip-worn sensor, where the magnitude of PA volume is expected to be substantially lower than when measured at wrist. Fourth, our harmonization mapping was estimated using generalized additive models (GAMs) and does not offer an easy, closed-form formula of the transformation. While such a formula could be obtained e.g. with polynomial models, the choice of GAMs allowed for thorough estimation of a relationship between AC and other measures in a more flexible way. Finally, our results are conditional on the data preprocessing methods we have chosen. However, we believe that the steps we performed are commonly done (we have cited studies who have previously used these) and are reasonable given the obtained data summary statistics and visual quality checks performed.

## Conclusions

In conclusion, our comparison of AC and MIMS, ENMO, MAD and AI allowed to show their high correlation to enhance comparability across past and future research. The derived harmonization mapping is freely available and provides a way to harmonize accelerometry data sets where summary measures were derived using different methods. Further research is warranted to test the validity of the mapping with different frequency data and body locations.

## Data Availability

The data used are a part of the National Institute on Aging's Baltimore Longitudinal Study of Aging (BLSA). National Institutes of Health (NIH) IRB approval is needed for a specific research proposal on these data.

## Declarations

### Ethics Approval and Consent to Participate

The study protocol was approved by the Internal Review Board of the Intramural Research Program of the National Institutes of Health. All participants provided written informed consent.

### Availability of Data and Materials

Data used in this manuscript were collected as part of the National Institute on Aging’s Baltimore Longitudinal Study of Aging (BLSA). National Institutes of Health IRB approval is needed for a specific research proposal to access these data. The project repository containing all code for data preprocessing and analysis is publicly available on GitHub (muschellij2/blsa_mims).

### Authors’ Contributions

Marta Karas implemented the final version of the analyses and took the lead in finalizing the manuscript. John Muschelli co-initiated the work, wrote the first draft of this manuscript and the analyses, and authored the SummarizedActigraphy R package. Ciprian M Crainiceanu provided scientific input and provided major edits to the manuscript draft. Jennifer A Schrack co-initiated the work, provided scientific input, and led the interpretation of results. Andrew Leroux, Jacek K Urbanek, Amal A Wanigatunga, and Jiawei Bai provided scientific input and helped with the interpretation of results. All authors provided critical feedback and helped shape the research, analysis, and manuscript.

## Abbreviations

AC: ActiGraph’s activity counts
AI: Activity Intensity
BLSA: Baltimore Longitudinal Study on Aging
ENMO: Euclidean Norm Minus One
MAD: mean amplitude deviation
MAPE: mean absolute percentage error
MIMS: Monitor-Independent Movement Summary
MPE: mean percentage error
PA: physical activity
TAC: total AC (total ActiGraph’s activity counts)

### Appendix A

#### Raw Accelerometry Data Quality Control

Three raw data quality check flags were adapted from a set of nine flags recently introduced by the release of the NHANES 2011-2012 wave protocol (NHANES 2011-2012 Data Documentation, 2020). The selected flags subset represents intuitive flags that are meant to “determine signal patterns that were unlikely to be a result of human movement” but are not aimed at identifying non-wear. To provide the flags definition, we denote a raw data observation as a vector **x**(t) = (x_1_(t), x_2_(t), x_3_(t)), where x_m_(t) is an acceleration measurement along axis m = 1,2,3 at time t.

First, large changes in acceleration values (“spikes”) were identified. Specifically, an observation **x**(t) was flagged if for any axis m = 1,2,3, x_m_(t) had an adjacent observation, x_m_(t − 1) or x_m_(t + 1), with an absolute difference greater than a threshold of 14.7 *g*. The 14.7 *g* threshold was adapted from NHANES protocol (11 *g*), as the NHANES devices had a dynamic range of 12 *g*, and the BLSA devices had a range of 16 *g*, so our threshold is proportional (11/12) to our data. Second, an observation **x**(t) was flagged if any axis measurement x_m_(t) occurred near the device maximum limit (here: 8 *g*, NHANES protocol: 6 *g*) with a tolerance margin (0.05 *g*), which translates to x_m_(t) being equal to or greater than 7.95 *g*. Third, an observation **x**(t) was flagged if any axis measurement x_m_(t) occurred near the device minimum limit (here: -8 *g*, NHANES protocol: -6 *g*), including a tolerance margin (0.05 *g*), and had same-axis adjacent observation also near the device minimum limit. These three flags were combined and the raw data observation **x**(t) was flagged as valid if it had none of the three flags; otherwise it was invalid.

### Appendix B

#### Open-source Summary Measures of Raw Accelerometry Data

The raw accelerometry data were used to derive a set of commonly used minute-level open-source summary measures: MIMS, ENMO, MAD, and AI. To provide the measures definition, we denote a raw data observation as a vector **x**(t) = (x_1_(t), x_2_(t), x_3_(t)), where x_m_(t) is an acceleration measurement along m = 1,2,3 axis collected at time t.

##### Monitor Independent Movement Summary (MIMS)

John et al. (2019) proposed Monitor-Independent Movement Summary unit (MIMS-unit). The MIMS-unit algorithm steps are conducted independently for each axis’ univariate acceleration signal x_m_(t), m = 1,2,3 until a final aggregation step. First, an input signal x_m_(t) is extrapolated to address a possible case when detected acceleration exceeds a sensor’s dynamic range; in this procedure, x_m_(t) is interpolated to 100 Hz, and then the extrapolation algorithm is applied to identify observations that hit the device limit (here: ± 8 *g*) and replace them with spline-interpolated points derived from the estimated extrapolation peak. The rest of the computations are done on this 100 Hz data. Second, a fourth-order Butterworth bandpass filter (0.2-5 Hz) is applied. Third, the interpolated, extrapolated, and filtered signal, 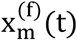, is aggregated within an epoch by computing area under curve via numerical integration; here, the epoch was set to 1 minute. Fourth, integrated values from each of the three axes are summed, yielding one value per epoch. Finally, the values less than or equal to 0.0001 * (epoch in seconds) * (sample rate after interpolation) (here: 0.0001 * 60 * 100 = 0.6) are truncated to zero.

The MIMS procedure may produce a negative value (−0.01), which indicates “the algorithm is unable to output a valid MIMS value for the given piece of the signal” (Tang et al., 2020). Negative MIMS output values were set to missing observations.

The algorithm implementation is provided in the MIMSunit R package (Tang et al., 2020). To compute MIMS, the package’s method mims_unit was used with its default values of internal parameters (consistent with the manuscript recommendations).

##### Euclidean Norm Minus One (ENMO)

Van Hees et al. (2013) proposed Euclidean Norm Minus One (ENMO) summary measure. ENMO calculation is based on Euclidean norm of (x_1_(t), x_2_(t), x_3_(t)),

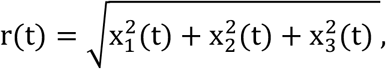

where negative values after subtracting are set to zero. Explicitly, the ENMO measure per epoch of H observations starting at time t_0_ is defined as 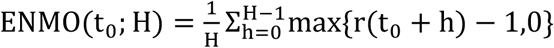

Here, H of size 60 * 80 = 4800 observations was used to match the number of observations in one minute with frequency of our raw accelerometry data. For each minute, t_0_ was set to be the time of the first observation within that minute.

Following the recommendations from van Hees et al. (2014), raw data calibration was performed for each participant separately before computing the ENMO measure. The g.calibrate method from the GGIR R package (van Hees et al., 2021) was used to estimate calibration values that were further used to center and scale the data accordingly. No other measures used the post-calibrated data.

##### Mean Amplitude Deviation (MAD)

Vähä-Ypyä et al. (2015) introduced Mean Amplitude Deviation (MAD) as a summary measure for accelerometry data. MAD measure per epoch of H observations starting at time t_0_ is defined as

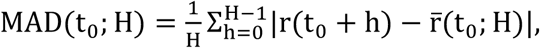

where 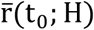 is defined as average Euclidean norm in the epoch, formally

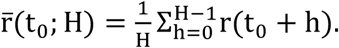

Here, H and t_0_ values were defined the same as for ENMO.

##### Activity Index (AI)

Bai et al. (2014) proposed the (unnormalized) Activity Index (AI) measure based on the combination of the three within-axis variance statistics. The variance of acceleration along m-th axis in the window of length Hstarting at time t_0_ is defined as

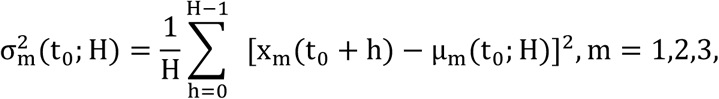

where μ_m_(t_0_; H) is axis-specific mean acceleration in the window, formally

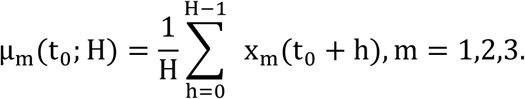

Then AI measure per epoch of H observations starting at time t_0_ is originally defined as

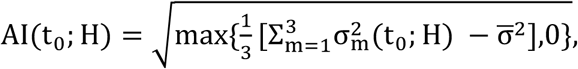

where 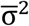 is the systematic noise variance calculated using the data collected during some non-moving period. In our work, 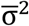 is not estimated and is set to zero in the above equation; hence, the AI formula used narrows down to

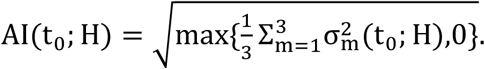

In computation of AI, first, a window H of size 1 * 80 = 80 was used to match the number of observations in one second with frequency of our raw accelerometry data, and t_0_ was set to be the time of the first observation within each second. Next, the per-second AI values were summed up within each minute so as the final outcome is defined at the minute level. This procedure is consistent with the recommendations given in Bai et al. (2016).

### Appendix C

#### Results

**Table C1.**
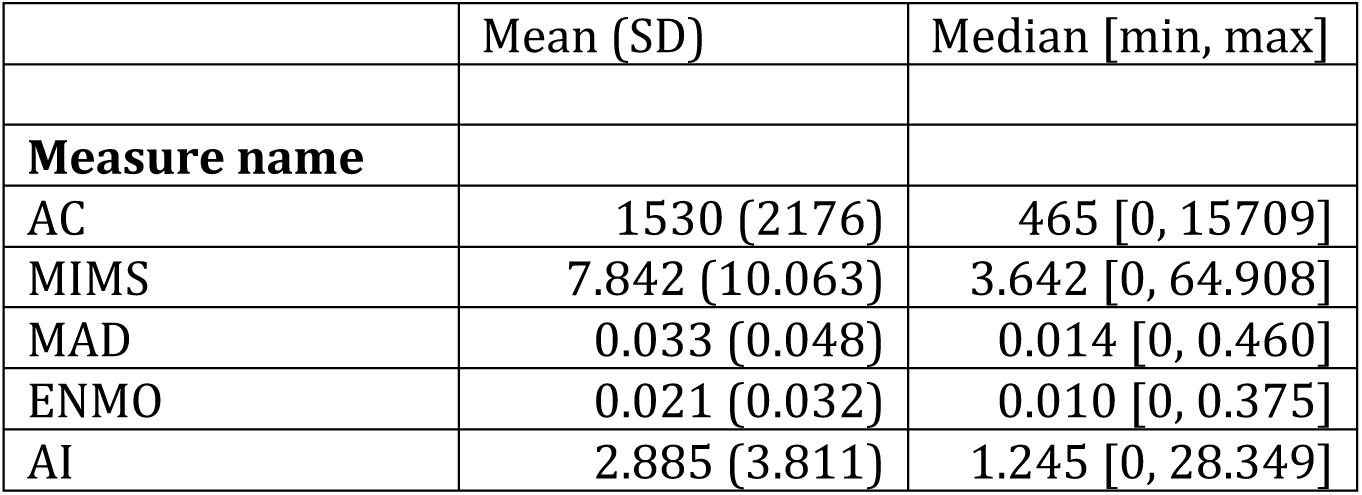
Summary of minute-level measures: AC, MIMS, ENMO, MAD, AI. Each value in the table is an aggregate -- mean, standard deviation, median, minimum and maximum -- of all participant-minutes. The summary was computed based on a data set after winsorization and invalid minutes data imputation.

**Table C2.** Summary of participant-specific mean percentage error (MPE) and mean absolute percentage error (MAPE) in estimating total activity counts (TAC).

|  | MPE | MAPE |
| --- | --- | --- |
|  | Mean (SD) | Mean (SD) |
| **Measure name** |  |  |
| MIMS | 0.2 (3.2) | 2.5 (2.4) |
| MAD | 4.6 (16.1) | 14.3 (10.3) |
| ENMO | -0.3 (13.3) | 11.3 (8.4) |
| AI | 0.3 (7.6) | 6.3 (5.1) |

**Figure C1.**
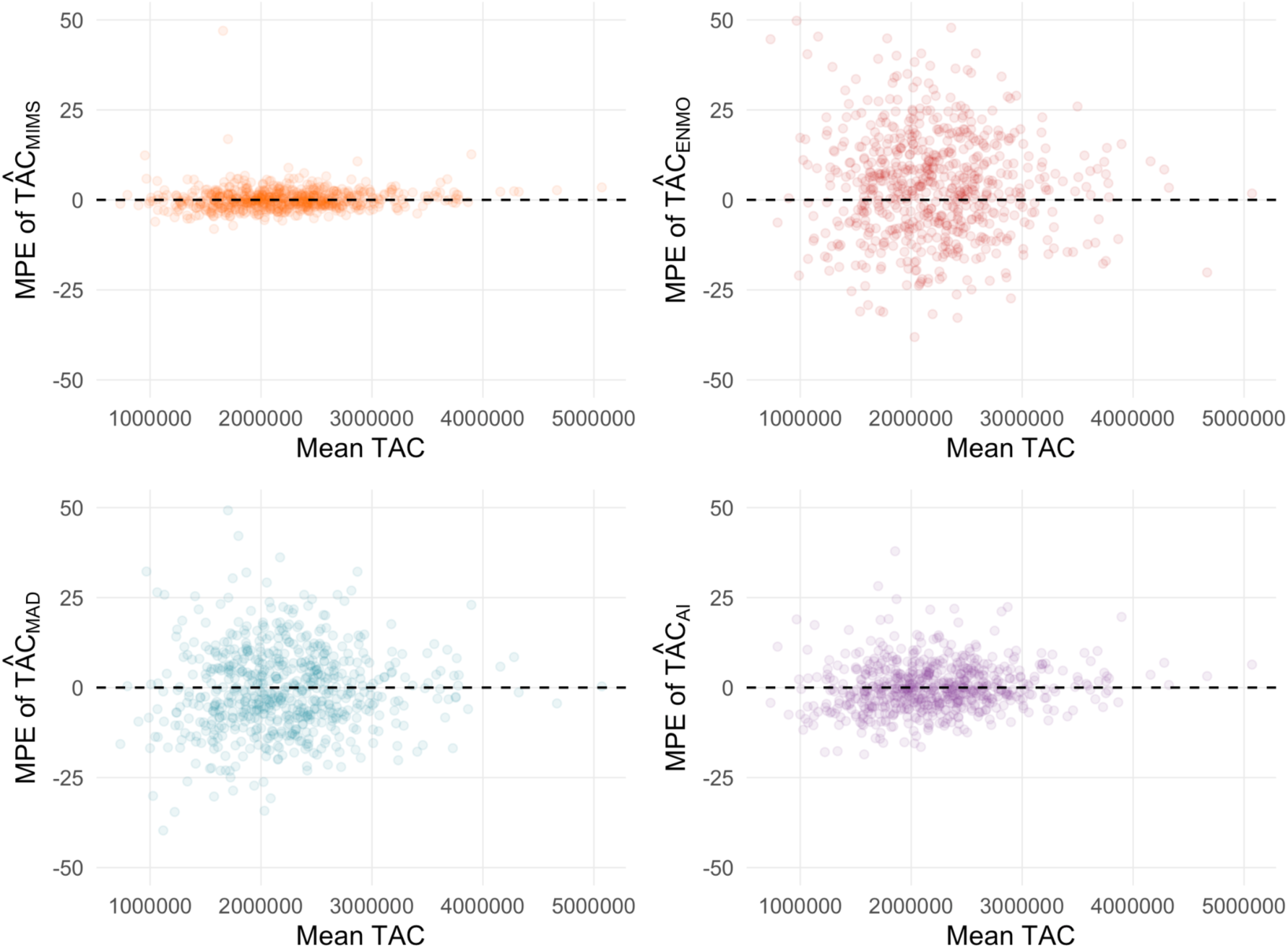
Participant-specific mean percentage error (MPE) in estimating total activity counts (TAC), arranged according to the participant’s average TAC. Each point represents one participant’s MPE.

**Table C3.** Summary of participant-specific accuracy, sensitivity and specificity of predicting whether a minute is active. The minute’s label was based on true AC and the prediction was based on 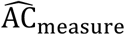. For each participant, performance metrics -- accuracy, sensitivity and specificity -- were computed across all participant’s minutes. Columns 3-5 show mean and standard deviation of participant-specific performance metrics.

|  |  | Accuracy | Sensitivity | Specificity |
| --- | --- | --- | --- | --- |
|  |  | Mean (SD) | Mean (SD) | Mean (SD) |
| Min.'s label<sup>a</sup> | Min.'s pred.<sup>b</sup> |  |  |  |
| AC > 1853 | $\widehat{AC}_{\text{MIMS}} > 1853$ | 0.981 (0.005) | 0.968 (0.012) | 0.986 (0.007) |
| AC > 1853 | $\widehat{AC}_{\text{ENMO}} > 1853$ | 0.904 (0.028) | 0.856 (0.071) | 0.921 (0.043) |
| AC > 1853 | $\widehat{AC}_{\text{MAD}} > 1853$ | 0.928 (0.021) | 0.879 (0.061) | 0.945 (0.028) |
| AC > 1853 | $\widehat{AC}_{\text{AI}} > 1853$ | 0.960 (0.012) | 0.933 (0.030) | 0.970 (0.018) |

<sup>a</sup>Min.s' label = condition defining a minute's true label in the classification task.
<sup>b</sup>Min.s' pred. = condition defining a minute's predicted label in the classification task.

